# Payments to key opinion leader physicians and drug sales of top pharmaceutical companies during the COVID-19 pandemic

**DOI:** 10.1101/2022.01.08.22268942

**Authors:** José Luis Sandoval, Alex Friedlaender, Alfredo Addeo, Glen J. Weiss

## Abstract

**Background:** The unprecedented context of the COVID-19 pandemic poses the opportunity to study several questions in circumstances that would probably not otherwise occur. We sought to determine the dynamics of pharmaceutical company drug sales revenue, market capitalization and payments to physicians during the pandemic, focusing on payments to so-called key opinion leaders (KOLs).

**Methods:** We analyzed the CMS Open Payments data of 15 top pharmaceutical company general payments to US physicians. We calculated total payments per year for all physicians, KOLs and 2018 KOLs in subsequent years. Drug-related fold changes in payments, drug revenues and company market capitation were calculated using Q1-2018 as reference. Yearly differences in payments, drug sales revenue and market capitalization were tested using generalized estimation equations (GEE). A double-sided p<0.05 was considered significant.

**Results:** The analyzed dataset comprised 8,563,872 payments to 382,779 physicians. In 2020, we observed a reduction in payments to physicians and KOLs compared to prior years. The total amount per KOL physician per company also decreased for each year for KOLs and the 2018 KOLs in the subsequent years. Payments per drug, but neither drug revenues nor pharmaceutical company market capitalization, followed a downward trend in 2020 compared to prior years. GEE analysis confirmed that, compared to 2018, the decrease in payments to KOLs overall and for the top drugs of each company was statistically significant. Yet, no significant differences in drug sales revenue and market capitalization was observed.

**Conclusions:** A substantial and significant reduction in payments to KOLs during the first fiscal year of the COVID-19 pandemic was not associated with a reduction in drug sales revenue of blockbuster drug products and the market capitalization of 15 top pharmaceutical companies. Overall, these findings suggest that a substantial part of pharmaceutical payments to KOLs do not appear to impact top drug sales revenues.

## Background

The unprecedented context of the COVID-19 pandemic poses the opportunity to study several questions in circumstances that would probably not otherwise occur.(1,2) The day-to-day marketing relationships between physicians and pharmaceutical companies were disrupted, with most medical conferences being cancelled or taking place virtually.(3)

## Objective

We sought to determine the dynamics of pharmaceutical company drug sales revenue, market capitalization and payments to physicians(4) during the pandemic, focusing on payments to so-called key opinion leaders (KOLs).

## Methods and Findings

We analyzed the CMS Open Payments data (https://www.cms.gov/openpayments) of 15 top pharmaceutical companies’ general payments to US physicians. Since the global reach of the pandemic was evident after March 2020, we considered payments from the second to fourth fiscal quarters of each year. Payments to non-physicians (e.g. dentists, chiropractors) and institutions were excluded. Royalties and ownership bonuses were also excluded. In addition, we excluded Celgene and Boehringer Ingelheim due to the company’s acquisition in 2019(5) and lack of detailed publicly available pharmaceutical sales revenue data, respectively. KOLs were defined as physicians in the top 1% of payments received per pharmaceutical company. We calculated total payments per year for all physicians, KOLs and 2018 KOLs in subsequent years. For KOLs, we determined annual payments per physician per company. We also determined total KOLs’ payments related to the top 5 drug products per company based on physician payments. Quarterly revenues records and average market capitalizations were collected from corporate financial reports. Currencies were converted to USD using the quarterly average exchange rate obtained from https://www.stlouisfed.org/.

Drug-related fold changes in payments, drug revenues and company market capitation were calculated using Q1-2018 as reference. For drug sales revenue analyses, we excluded drugs lacking data for more than one fiscal quarter. For four drugs, we used Q2-2018 as reference due to Q1-2018 missing data. Individual physician payments related to multiple drugs were divided by the number of drugs identified for that payment. Yearly differences in payments, drug sales revenue and market capitalization were tested using generalized estimation equations (GEE). Data were analyzed with R (v4.1.0) using the *geepack* package. A double-sided p<0.05 was considered significant.

The analyzed dataset comprised 8,563,872 payments to 382,779 physicians. In 2020, we observed a reduction in payments to physicians and KOLs compared to prior years (**Figure 1A**). The total amount per KOL physician per company also decreased for each year’s KOLs and the 2018 KOLs in the subsequent years (**Figure 1B**). Total payments related to the top 5 drugs of each company also decreased during 2020 (**Figure 1C**). Payments per drug also followed a downward trend in 2020 compared to prior years (**Figure 1D**). However, the drug revenues of the analyzed drugs did not follow this negative trend (**Figure 2A-B**). Consequently, pharmaceutical companies’ market capitalization did not reflect the downward trend in physician payments (**Figure 2C**). GEE analysis confirmed that, compared to 2018, the decrease in payments to KOLs overall and for the top drugs of each company was statistically significant (**Figure 1E**). Yet, no significant differences in drug sales revenue and market capitalization was observed (**Figure 2D**).

**Figure 1.**
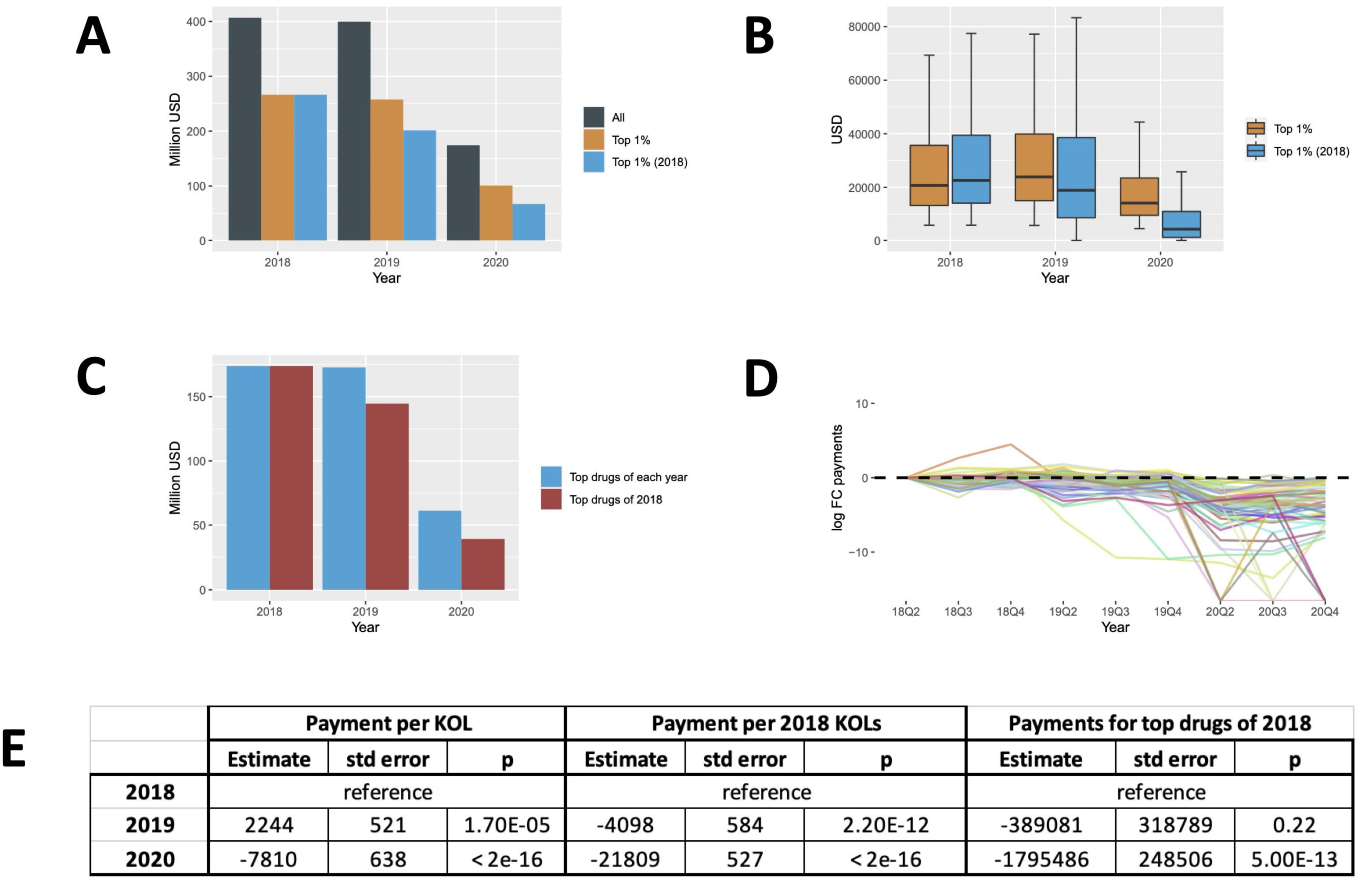
Payments to physicians by the selected 15 pharmaceutical companies during 2020 and the preceding years. A) total payments to physicians (all, top 1% of each year and 2018); B) payments per physician per company (top 1% of each year and 2018); C) total payments related to each company’s top 5 drugs of each year and 2018; D) Fold change of payments related to each company’s 2018 top 5 drugs; E) Results of GEE models testing for yearly differences of physician payments during the analyzed periods (2018,2019,2020).

**Figure 2.**
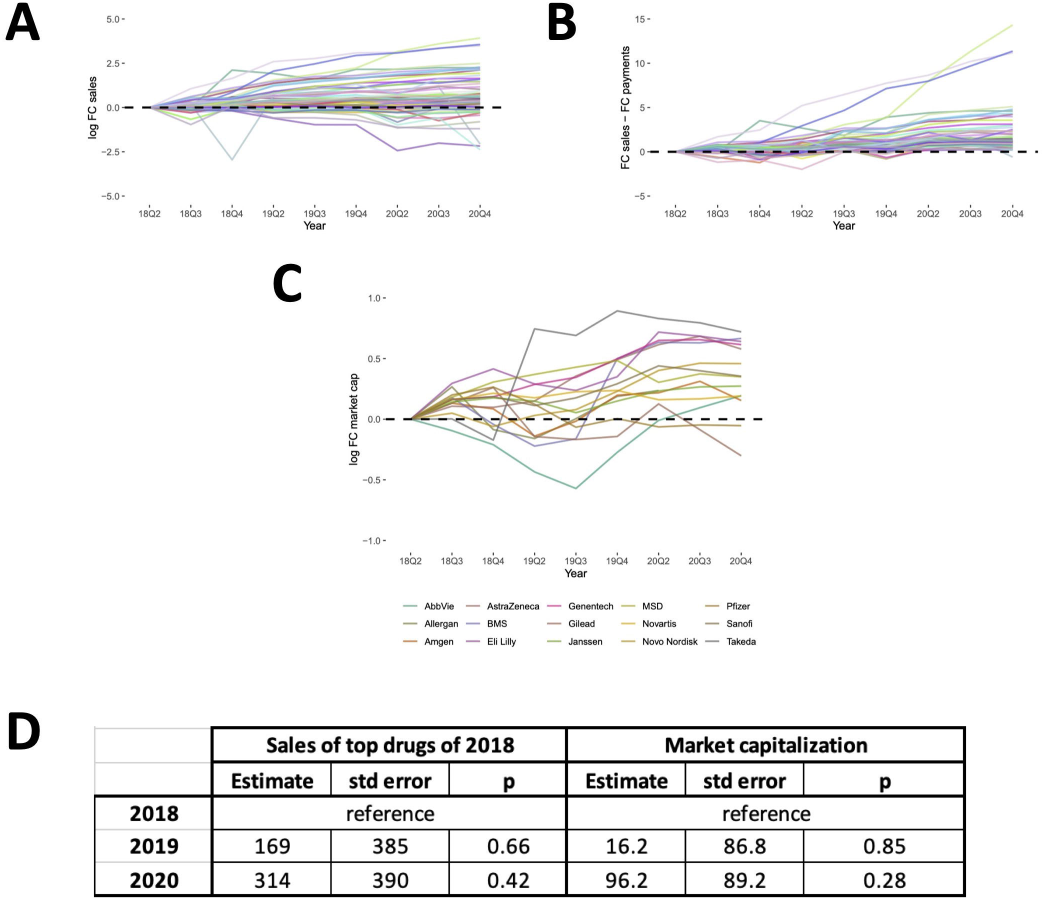
Sales revenue and market capitalization. A) Fold change of sales revenue related to each company’s 2018 top 5 drugs; B) Differential between fold changes of sales revenue and associated physician payments; C) Fold change of selected companies market capitalization; D) Results of GEE models testing for yearly differences of drug sales revenue and market capitalization during the analyzed periods (2018,2019,2020).

## Discussion

A substantial and significant reduction in payments to KOLs during the first fiscal year of the COVID-19 pandemic was not associated with a reduction in drug sales revenue of blockbuster drug products and the market capitalization of 15 top pharmaceutical companies. Overall, these findings suggest that a substantial part of pharmaceutical payments to KOLs do not appear to impact top drug sales revenues.

## Data Availability

All data produced in the present study are available upon reasonable request to the authors.

